# Long-Term Effect of Early-Life Arsenic Exposure on Morning Plasma Cortisol in Adults from Antofagasta, Chile

**DOI:** 10.1101/2025.09.01.25334887

**Authors:** Rosemarie de la Rosa, Craig Steinmaus, Anthony Nardone, Amanda Keller, Johanna Acevedo, Catterina Ferreccio, Martyn T. Smith, Fenna C. M. Sillé

**Author notes:** **Corresponding Author**: Rosemarie de la Rosa, PhD, MPH. 2121 Berkeley Way, Room 5302, University of California Berkeley, California 94720-7360.

## Abstract

Over 100,000 people were exposed to arsenic-contaminated drinking water in Antofagasta, Chile from 1958-1970. Individuals born during this high exposure period have elevated rates of cancer, lung and cardiovascular disease, and hypertension. However, the mechanisms of long-term arsenic toxicity remain unclear. We investigated whether early-life arsenic exposure was associated with altered glucocorticoid levels in adulthood. This study included 114 individuals born in Antofagasta during the high exposure period and 118 individuals born elsewhere.

Arsenic exposure metrics were constructed based on residential histories and included: concentration at birth, peak and highest 5-year average between ages 0-10 years, and highest lifetime 5-year average, and lifetime cumulative exposure. Morning plasma cortisol concentrations were measured using a cell-based bioassay. Individuals in the highest quartile of highest lifetime 5-year average of arsenic exposure had approximately 11% lower mean log cortisol levels than those in the lowest quartile of exposure (β = -0.116; 95% CI: -0.229, -0.003). In sex-stratified analyses, associations were stronger among females. For example, females in the highest quartile of cumulative exposure had 22.0% lower cortisol levels compared to those in the lowest quartile (β = -0.248; 95% CI: -0.444, -0.053) and the test for interaction by sex was statistically significant (p = 0.036). This study is the first to show that early-life arsenic exposure may have lasting effects on cortisol. These findings highlight endocrine disruption as a mechanism contributing to long-term health effects of early arsenic exposure.

## 1. Introduction

An estimated 220 million people worldwide are exposed to arsenic-contaminated drinking water at levels that exceed the World Health Organization (WHO) provisional guideline of 10 µg/L (Podgorski and Berg, 2020). This widespread exposure to arsenic poses a major global public health concern. Inorganic arsenic is a Group 1 human carcinogen and has been associated with a wide range of health effects, including skin lesions, cardiovascular disease, hypertension, diabetes, and adverse developmental outcomes (National Research Council, 2013; Naujokas et al., 2013). Furthermore, the developing fetus and children are particularly vulnerable to arsenic exposure, as early-life exposure can have long-term effects that increase the risk of chronic diseases in adulthood (Smeester and Fry, 2018). Therefore, there is an important need to better understand how early-life arsenic exposure leads to long-term health outcomes.

Northern Chile presents a unique and important opportunity to investigate the long-term health effects of arsenic exposure from drinking water. This geographic area contains the Atacama Desert, which is one of the driest habitable places on earth. Water sources are limited because of the arid climate and each city essentially has its own single water supply. Records on arsenic concentrations in all major water sources in the area are available for past decades (Ferreccio et al., 2000). The relatively few water sources and availability of good historical records allows for the construction of accurate lifetime arsenic exposure estimates for individuals, from birth through adulthood, simply by knowing the cities in which they have lived. Another unique feature of this region in Chile is that one of the largest cities, Antofagasta, had a well-documented distinct period of exceptionally high arsenic exposure. This exposure began in 1958 when rapid population growth led to supplementation of the city’s water supply with rivers containing arsenic concentrations of 860 μg/L and ended in 1970 with the installation of an arsenic removal treatment plant (Ruffino et al., 2022). As a result, more than 100,000 people were exposed to arsenic concentrations that exceeded the current WHO arsenic standard by greater than 80-fold for a period of 12 years. This exposure scenario, with its distinct start and stop, large population, and good exposure records provides a unique opportunity to investigate the long-term consequences of arsenic exposure. Ecological studies conducted in this region of Chile observed that adults exposed to high levels of arsenic-contaminated drinking water *in utero* and during early childhood had higher mortality rates from bronchiectasis (Smith et al., 2006), acute myocardial infarction (Yuan et al., 2007), and cancer (Yuan et al., 2010; Smith et al., 2012) compared to those who were lesser exposed or exposed only as adult. These associations persisted 30-40 years after the high exposure period ended (Roh et al., 2018). In addition to mortality, follow-up studies in this same population observed a higher incidence of lung and bladder cancer (Steinmaus et al., 2014), impaired lung-function (Dauphiné et al., 2011; Steinmaus et al., 2016) and increased prevalence of hypertension (Hall et al., 2017) among adults with high *in utero* and early-life arsenic exposure. Collectively, these studies provide strong evidence that early-life arsenic exposure induces persistent alterations that promote disease into adulthood. Therefore, understanding biological mechanisms underlying these persistent effects is crucial to inform public health strategies to mitigate the long-term health effects of early-life arsenic exposure.

The biological processes underlying persistent effects of early-life arsenic exposure are not well understood. Several mechanisms have been proposed, including altered epigenetic reprogramming, immune modulation, and oxidative stress (Bailey et al., 2016). These mechanisms may also contribute to the endocrine-disrupting effects of arsenic on the hypothalamic-pituitary-adrenal (HPA) axis, a highly conserved neuroendocrine system that regulates the body’s response to stressors and maintains metabolic and immune homeostasis through the secretion of glucocorticoids (cortisol in humans and corticosterone in rodents) (Sun et al., 2016). Animal studies have demonstrated that prenatal arsenic exposure induces long-term HPA axis dysfunction that persists into adulthood, characterized by elevated basal corticosteroid levels, blunted stress reactivity, and impaired negative feedback (Martinez et al., 2008; Goggin et al., 2012). Prenatal arsenic exposure has also been shown to alter expression and protein levels of key signaling genes involved in HPA axis regulation, including the glucocorticoid receptor (*NR3C1*) and 11β-hydroxysteroid dehydrogenase enzymes (*11β-HSD1 and 11β-HSD2*) (Caldwell et al., 2015; Goggin et al., 2012). Despite strong mechanistic evidence from animal models, epidemiological studies examining the effects of arsenic exposure on plasma cortisol (the human equivalent of corticosterone) are limited and have yielded inconsistent findings. In a study of 267 women aged 22-45 years from West Bengal, India, those currently exposed to drinking water containing 11-50 μg/L of arsenic had two-fold higher plasma cortisol levels than women exposed to <10 μg/L arsenic (Sinha et al., 2014). While results from this study were consistent with those from animal studies, other studies have observed inverse associations between arsenic exposure and cortisol levels. A cohort study of 168 mother-child dyads from Arica, Chile found that among higher income families greater second-trimester urinary arsenic concentrations in mothers were associated with lower salivary cortisol in their infants 18-24 months after delivery (Salgado et al., 2019). Similarly, plasma arsenic concentrations were inversely correlated with plasma cortisol levels among a convenience sample of male runners (Alves et al., 2020). However, null associations have also been reported, with no difference in plasma or urinary cortisol concentrations observed with greater arsenic exposure (Chen et al., 2024; Li et al., 2020). Despite these mixed findings, the presence of either hypercortisolism (e.g. Cushing’s syndrome) or hypocortisolism could indicate perturbed HPA axis regulation, which is known to result in adverse clinical outcomes (Reincke and Fleseriu, 2023; Vaidya et al., 2025). These inconsistent findings may reflect variability in exposure characteristics (e.g., timing, concentrations, and sources), as well as differences in how both exposure and cortisol were measured, or other demographic factors. It should be noted that all but one study was cross-sectional and that study examined the relationship between prenatal arsenic exposure and cortisol only during infancy (Salgado et al., 2019). Additional longitudinal studies are needed to determine whether early-life arsenic exposure influences cortisol regulation beyond this developmental period and across the lifecourse. This study is the first to assess whether early-life arsenic exposure has a persistent effect on plasma cortisol concentrations into adulthood in humans.

The effects of early-life arsenic exposure on the HPA axis also appear to differ by sex (Allan et al., 2015; Solomon et al., 2020). In male mice, prenatal exposure reduced glucocorticoid receptor and Hsd11b1 protein levels and increased FKBP5 acetylation and expression. Female exposed mice appeared to be resistant to these biological changes, possibly due to greater antioxidant capacity. Further research is needed to better understand the molecular and sex-specific effects of early-life arsenic exposure on the HPA axis in humans.

Here, we investigated associations between early-life and lifetime arsenic exposure with morning plasma cortisol concentrations in adulthood. This study included 233 adults currently living in Antofagasta, with 114 individuals born in Antofagasta during the high exposure period and 118 individuals who were born elsewhere in Chile with lower levels of exposure. Because early-life arsenic exposure has been shown to have sex-specific effects on HPA axis activity in animal studies, we also explored sex differences in the association between arsenic exposure and plasma cortisol. Another unique aspect of this study is the use of a rapid, low-cost cell-based bioassay to measure differences in plasma cortisol concentrations between individuals with high versus low arsenic exposure. This study population provides a rare opportunity to investigate the long-term consequence of early-life and lifetime arsenic exposure on cortisol levels in adulthood.

## 2. Methods

### 2.1 Study Participants

Participants were a convenience sample of employees from the Antofagasta Hospital and University of Antofagasta as well as participants from general recruitment across Antofagasta in Chile. Recruitment occurred during two time periods: November 2013-January 2014 and May-July 2017. The study included 114 participants who were born in Antofagasta during the high exposure period (1958-1970) and who still resided in Antofagasta at the time of recruitment along with a comparison group of 118 participants that were born elsewhere and moved to Antofagasta after the high exposure period. Out of the 250 individuals initially recruited for the study, only those born during the high exposure period (1958-1970) were included in this analysis (n=233). Consistent with the 1958-1970 exposure window, participant ages at recruitment were 43-55 years for the 2013 study and 47-59 years for the 2017 study. We also excluded one participant that had plasma cortisol levels below the assay’s limit of detection (described in section 2.3), resulting in a final sample of 232 participants. When comparing demographics between the 2013 and 2017 recruitment periods, the latter tended to be older (median age 48 vs 54 years), included more females (35.7% vs. 50.0%), ever smokers (66.7% vs 55.4%), and had less participants who completed secondary school (88.1% vs 75.7%) than in 2013 (Supplemental Table 1). Arsenic exposure levels were also greater among participants in the 2017 study. Since there were significant differences in sociodemographic characteristics and arsenic exposure between participants recruited in 2013 and 2017, recruitment period was included as a covariate in our primary analyses. Stratified analyses by recruitment period were also conducted as a sensitivity analysis.

Exclusion criteria for the study included antibiotic use in the past 3 months, use of enemas or laxatives more than once per month, or use of steroids or immunosuppressants. Data and sample collection for this study were approved by institutional review boards at the University of California, Berkeley and the School of Medicine at the Pontificia Universidad Católica de Chile.

### 2.2 Arsenic Exposure

We collected a detailed lifetime residential history from each participant using a standardized questionnaire. Participants were asked to provide all residences where they lived for ≥ 6 months, the water source at each residence (e.g. bottled water, tap), and their typical daily water intake currently as well as 20 and 40 years ago. We then linked each participant’s city of residence to annual arsenic water measurements obtained from government agencies, water companies, and other sources that were available for all large cities and towns in Chile (Ferreccio et al., 2000). While arsenic levels outside of Antofagasta were generally below 10 µg/L (Smith et al., 2018), it is important to note that some nearby towns also had elevated arsenic levels in their drinking water supply. While not as high as Antofagasta, arsenic concentrations at birth for 47 of 118 participants born elsewhere between 1958-1970 still exceeded Chile’s national drinking water standard of 50 µg/L (this standard was later reduced to 10 µg/L in 2005). Therefore, we also calculated several arsenic exposure metrics for each participant to assess both the intensity and cumulative nature of exposure. These individual-level arsenic exposure metrics included: born in Antofagasta during the high exposure period (1958-1970) as a binary variable; arsenic concentration in drinking water in their birth city; peak exposure during 0-10 years of life captured the highest single annual concentration during early-life; and highest 5-year average exposure during 0-10 years of life to assess sustained high exposure during early-life. The 0–10 year age range was selected because most participants’ cumulative arsenic exposure occurred during their first decade of life (Supplemental Figure 2). Lifetime highest 5-year average exposure was also estimated as the highest annual concentration averaged over any contiguous 5- year period. Lastly, lifetime cumulative arsenic exposure was calculated for each participant by summing yearly concentrations from birth until the time of sample collection.

### 2.3 Plasma Cortisol Equivalent Concentrations

A single fasted blood sample was collected in EDTA tubes from each willing participant and time of sample collection was recorded by research personnel. The average time of blood draw was 9:00 AM and ranged from 6:10AM to 12:35PM. All samples were processed within 8 hours upon collection and plasma aliquots were frozen at -80°C for 2-8 weeks before being transported on dry ice to the University of California, Berkeley where they were stored until analysis. Cortisol assays were performed in two separate batches. Samples collected during the 2013 recruitment period were performed on May 19, 2016 and those from the 2017 recruitment period were performed on February 10, 2018.

Plasma cortisol equivalent concentration values were obtained using the 231GRE cell-based bioassay developed by our research group (de la Rosa et al., 2021). We previously demonstrated high concordance correlation (r_c_=0.92) between this assay and cortisol enzyme-linked immunosorbent assay measurements in plasma collected from 12 healthy individuals. Briefly, the 231GRE cell line was generated by stably transfecting MDA-MB-231 cells with a luciferase reporter gene driven by three simple glucocorticoid-response elements. These cells were maintained at 37°C with 5% CO_2_ using Dulbecco’s Modified Eagle Medium (DMEM; Gibco) containing 10% fetal bovine serum (FBS; Atlanta Biologicals). One week prior to experiments, cells were cultured using phenol red-free DMEM (Hyclone) with 10% charcoal-dextran FBS (Atlanta Biological) to reduce the concentration of hormones present in media. Hormone-stripped cells were seeded at 2.7×10^4^ cells/well in white 96-well plates and incubated at 37°C overnight to allow cells to attach. The next day, media in quadruplicate wells was replaced with 100µL of media containing either a range of cortisol standards (0, 1.56, 3.13, 6.25, 12.5, 25 nM) or human plasma samples diluted 1:20 in hormone-depleted media. Cells were treated for 24 hours at 37°C before rinsing wells with PBS and adding 1x cell lysis buffer (Promega). Luminescence was quantified as relative light units (RLUs), which are proportional to the degree of luciferase reporter activity, using a Berthold Centro XS3 LB 960 microplate luminometer and an automatic injector for Luciferase Assay Reagent (Promega). Duplicate cortisol standards (0-25nM in 0.1% DMSO) and a common reference plasma sample (diluted 1:20 run in quadruplicate) were included on every plate.

RLUs reflect the total amount of glucocorticogenic compounds present in media containing diluted plasma samples. Wells with RLUs below the lowest cortisol standard on the plate (1.56nM) were exclude from the analysis to avoid bias from imputing values below this concentration. This resulted in one participant being dropped from the analysis. Sample RLUs were converted to cortisol equivalent concentrations by interpolating values from standard curves on each plate that were fit with an inverse weighted 4-parameter logistic regression model. The standard curves for cortisol on each plate fit well with a median R^2^ value of 0.994 (IQR: 0.991, 0.997) across all 26 plates. To minimize plate-to-plate variability and account for batch effects from running the 2013 and 2017 studies separately, cortisol equivalent concentration values were normalized using a common reference plasma sample included on each plate. Estimated cortisol concentrations for each well on a plate were divided by the median concentration of the reference sample on its respective plate and then multiplied by the overall median concentration of the reference sample across all plates. Plate (random) and batch (fixed) effects were assessed both before and after normalization with the reference sample. After normalization, neither effect remained statistically significant. These normalized cortisol equivalent concentration values were then averaged for each participant and multiplied by 20 to account for the plasma dilution factor. Assay CVs for all participant samples ranged from 0.6% to 23.8% with a mean of 6.3%.

### 2.4. Other Covariate Data

Covariates were selected based on existing literature examining associations between arsenic exposure or other heavy metals and cortisol levels (Choi et al., 2022; Gump et al., 2008; Pérez-Cadahía et al., 2008; Salgado et al., 2019; Souza-Talarico et al., 2017). Participants age was collected as a continuous variable measured in years. Sex (male vs. female), smoking status (ever vs. never), and highest education or grade achieved (< secondary school vs. ≥ secondary school) were assessed via questionnaire.

### 2.5 Statistical Analysis

Sociodemographic characteristics were compared between participants born in Antofagasta and those born elsewhere using Fisher’s exact test for categorical variables and Wilcoxon rank sum test for continuous variables. Plasma cortisol equivalent concentrations were log-transformed since its distribution was right-skewed. Unadjusted linear regression analyses were used to evaluate univariate associations between sociodemographic factors and plasma cortisol equivalent concentrations. Multivariable regression models were used to estimate mean differences in plasma cortisol equivalent concentration (dependent variable) by arsenic exposure group (independent variable). Being born in Antofagasta was treated as a binary variable (no/yes). Given the exposure range and non-linear relationship with cortisol (Salgado et al., 2019), arsenic concentration at birth was analyzed by tertiles with the lowest concentration group serving as the reference. The remaining arsenic exposure metrics were assessed using quantiles. Final models were adjusted for age, sex, smoking status, education level, time of blood sample collection, and study recruitment period (2013 vs. 2017). Blood sample collection time was included as hours since 6AM and modeled as a spline with 3 degrees of freedom to account for the diurnal rhythm of cortisol (Supplemental Figure 1). Given evidence from animal studies demonstrating sex-specific effects (Allan et al., 2015; Caldwell et al., 2015; Solomon et al., 2020), analyses between arsenic exposure groups and log-transformed cortisol equivalent concentrations were stratified by sex. A multiplicative interaction term between arsenic exposure and sex was also included to test for potential effect modification. Sensitivity analyses were conducted to rule out potential confounding by stratifying analyses by median age (52 years) and recruitment period. Percent differences in cortisol concentrations were calculated as [e^β^ – 1] × 100, where β is the coefficient from log-transformed models. All p-values were two-sided and a value <0.05 was considered statistically significant. Statistical analyses were conducted using R software, version 4.3.1.

## 3. Results

Of the total 232 study participants, 114 (49.1%) were born in Antofagasta during the high arsenic exposure period and 118 (50.9%) were born elsewhere. **Table 1** summarizes the sociodemographic characteristics for all participants as well as stratified by birth location (Antofagasta vs. elsewhere). Participants born in Antofagasta were comparable in age (median: 52 vs 51 years) and sex (54.4% vs 55.9% male) to those born elsewhere. The two groups were also similar in the proportion of participants from each recruitment period and their blood collection times. However, there were slightly fewer individuals who reported ever smoking (52.6% vs 66.1%) and completing secondary school (75.4% vs 84.7%) among participants born in Antofagasta compared to the those born elsewhere. All arsenic exposure metrics, except for lifetime average after age 20, were higher among participants born in Antofagasta. The median concentration of arsenic in drinking water at birth was 860 μg/L for participants born in Antofagasta and 6.2 μg/L for those born elsewhere. In contrast, lifetime average concentration levels after age 20 were comparable between the two exposure groups (median: 26 vs 24 μg/L).

**Table 1.**
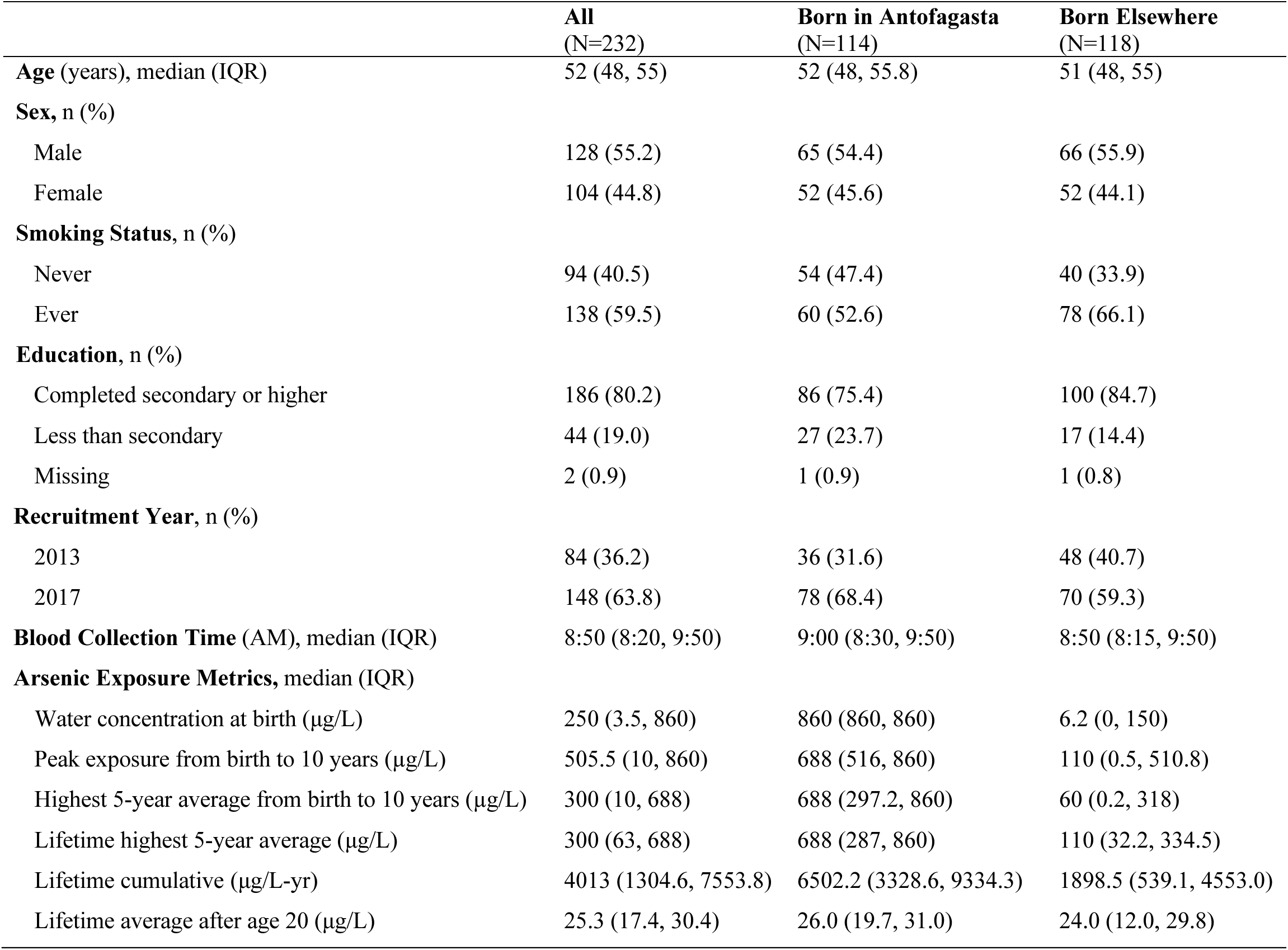
Characteristics of study population.

|  | **All**<br>(N=232) | **Born in Antofagasta**<br>(N=114) | **Born Elsewhere**<br>(N=118) |
| --- | --- | --- | --- |
| **Age** (years), median (IQR) | 52 (48, 55) | 52 (48, 55.8) | 51 (48, 55) |
| **Sex**, n (%) |  |  |  |
| Male | 128 (55.2) | 65 (54.4) | 66 (55.9) |
| Female | 104 (44.8) | 52 (45.6) | 52 (44.1) |
| **Smoking Status**, n (%) |  |  |  |
| Never | 94 (40.5) | 54 (47.4) | 40 (33.9) |
| Ever | 138 (59.5) | 60 (52.6) | 78 (66.1) |
| **Education**, n (%) |  |  |  |
| Completed secondary or higher | 186 (80.2) | 86 (75.4) | 100 (84.7) |
| Less than secondary | 44 (19.0) | 27 (23.7) | 17 (14.4) |
| Missing | 2 (0.9) | 1 (0.9) | 1 (0.8) |
| **Recruitment Year**, n (%) |  |  |  |
| 2013 | 84 (36.2) | 36 (31.6) | 48 (40.7) |
| 2017 | 148 (63.8) | 78 (68.4) | 70 (59.3) |
| **Blood Collection Time** (AM), median (IQR) | 8:50 (8:20, 9:50) | 9:00 (8:30, 9:50) | 8:50 (8:15, 9:50) |
| **Arsenic Exposure Metrics**, median (IQR) |  |  |  |
| Water concentration at birth (µg/L) | 250 (3.5, 860) | 860 (860, 860) | 6.2 (0, 150) |
| Peak exposure from birth to 10 years (µg/L) | 505.5 (10, 860) | 688 (516, 860) | 110 (0.5, 510.8) |
| Highest 5-year average from birth to 10 years (µg/L) | 300 (10, 688) | 688 (297.2, 860) | 60 (0.2, 318) |
| Lifetime highest 5-year average (µg/L) | 300 (63, 688) | 688 (287, 860) | 110 (32.2, 334.5) |
| Lifetime cumulative (µg/L-yr) | 4013 (1304.6, 7553.8) | 6502.2 (3328.6, 9334.3) | 1898.5 (539.1, 4553.0) |
| Lifetime average after age 20 (µg/L) | 25.3 (17.4, 30.4) | 26.0 (19.7, 31.0) | 24.0 (12.0, 29.8) |

**Table 2** lists associations between sociodemographic factors and log-transformed morning plasma cortisol concentration values. The median log cortisol concentration among participants was 5.1nM (range: 4.3-6.0nM). Blood collection time was inversely associated with cortisol concentrations, meaning values declined the later the blood was collected. Mean plasma cortisol concentrations were ∼15% lower among females compared to males (ß=-0.162; 95% CI: -0.239, -0.085). Cortisol concentrations did not differ by age, education, smoking status, or year of recruitment.

**Table 2.** Associations between sociodemographic factors and plasma cortisol equivalent concentrations.

| Variable | ln(Cortisol) (nM),<br>Median (IQR) | $\beta$ (95% CI) |
| --- | --- | --- |
| All | 5.11 (4.88, 5.34) |  |
| **Continuous** |  |  |
| Age (years) |  | -0.005 (-0.014, 0.004) |
| Blood Collection Time (hours since 6AM) |  | -0.070 (-0.106, -0.036) |
| **Categorical** |  |  |
| Sex |  |  |
| Male | 5.20 (4.98, 5.38) | Ref |
| Female | 5.01 (4.82, 5.24) | -0.162 (-0.239, -0.085) |
| Smoking Status |  |  |
| Never | 5.07 (4.84, 5.28) | Ref |
| Ever | 5.14 (4.92, 5.37) | 0.067 (-0.013, 0.148) |
| Education |  |  |
| Completed secondary or higher | 5.11 (4.88, 5.31) | Ref |
| Less than secondary | 5.13 (4.86, 5.37) | 0.003 (-0.098, 0.104) |
| Recruitment Year |  |  |
| 2013 | 5.17 (4.93, 5.37) | Ref |
| 2017 | 5.07 (4.86, 5.29) | -0.067 (-0.149, 0.015) |

Associations between arsenic exposure and morning plasma cortisol concentrations were examined using multivariable linear regression (**Table 3**). In unadjusted models, cortisol concentrations did not differ between individuals born in Antofagasta and those born elsewhere. Cortisol concentrations were similar between the highest and lowest tertile of arsenic concentration at birth. For peak arsenic exposure and highest 5-year average exposure during ages 0–10 years, individuals in the highest exposure quartile had approximately 11% lower mean cortisol concentrations compared to those in the lowest exposure quartile (ß _peak exposure 0-10y=860 μg/L_ = -0.117; 95% CI: -0.225, -0.010; ß _highest 5-year average 0-10y>688_ = -0.115; 95% CI: -0.227, -0.004).

**Table 3.**
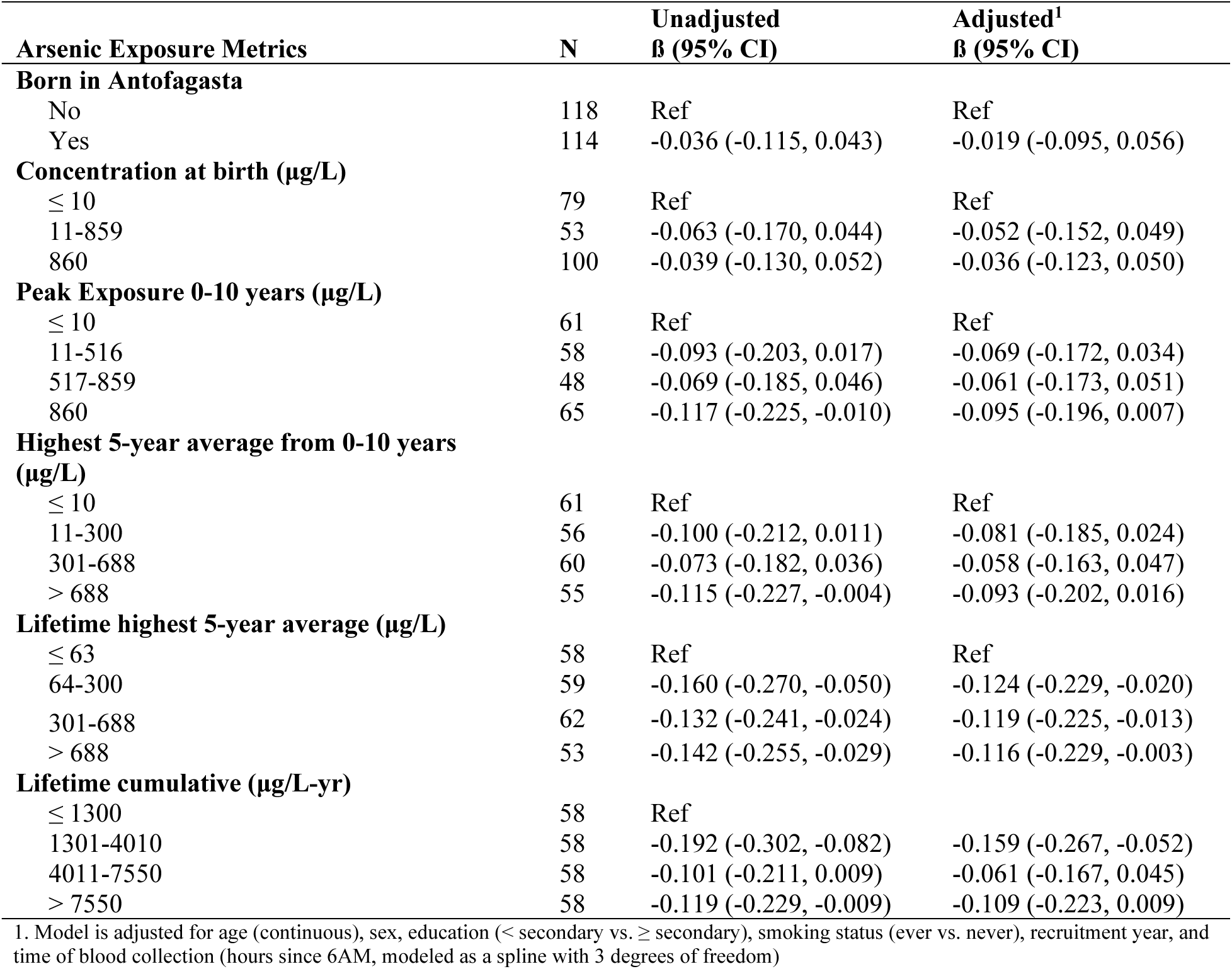
Associations between arsenic exposure metrics and plasma cortisol equivalent concentrations.

Cortisol concentrations were lower across all quartiles of highest 5-year average exposure above the lowest quartile (≤ 63 μg/L). For cumulative lifetime arsenic exposure, lower cortisol concentrations were observed for the second (ß = -0.192; 95% CI: -0.302, -0.082) and fourth (ß = -0.119; 95% CI: -0.229, -0.009) quartiles relative to the first exposure quartile. Adjusting models for covariates modestly attenuated effect sizes. Consequently, only associations with lifetime highest 5-year average exposure and the second quartile of cumulative lifetime exposure remained statistically significant after covariate adjustment.

Sex-stratified analyses were also conducted to assess if associations differed between males and female participants (Figure 1, Supplemental Table 2). Higher arsenic exposure was associated with lower plasma cortisol equivalent concentrations among both males and females. However, these effects were consistently stronger and only reached statistical significance among female participants. Specifically, females with a peak arsenic exposure of 860 μg/L between ages 0-10 years had mean cortisol levels that were 16.8% lower compared to participants whose highest level of exposure was ≤ 10 μg/L before age 10 (ß = -0.184; 95% CI: - 0.356, -0.012). Among males, this association was not statistically significant (ß = -0.043; 95% CI: -0.171, 0.086). We also observed that females in the second (ß = -0.196; 95% CI: -0.367, - 0.026) and fourth (ß = -0.200; 95% CI: -0.387, -0.014) quartiles of lifetime 5-year average arsenic exposure had approximately 18% lower mean cortisol concentrations than female participants in the first exposure quartile. The second (ß = -0.265; 95% CI: -0.441, -0.090) and fourth (ß = -0.248; 95% CI: -0.444, -0.053) quartiles of lifetime cumulative exposure were associated with 23.3% and 22.0% lower cortisol concentrations, respectively, than females in the first quartile. No statistically significant differences were observed among males for these exposure measures. When testing for effect modification by sex, a statistically significant interaction was observed with lifetime cumulative exposure (p-interaction=0.036), but not for any of the other exposure measures. These results demonstrate that the effect of lifetime cumulative arsenic exposure on cortisol concentrations differed between males and females, with a stronger association observed among female participants.

**Figure 1.**
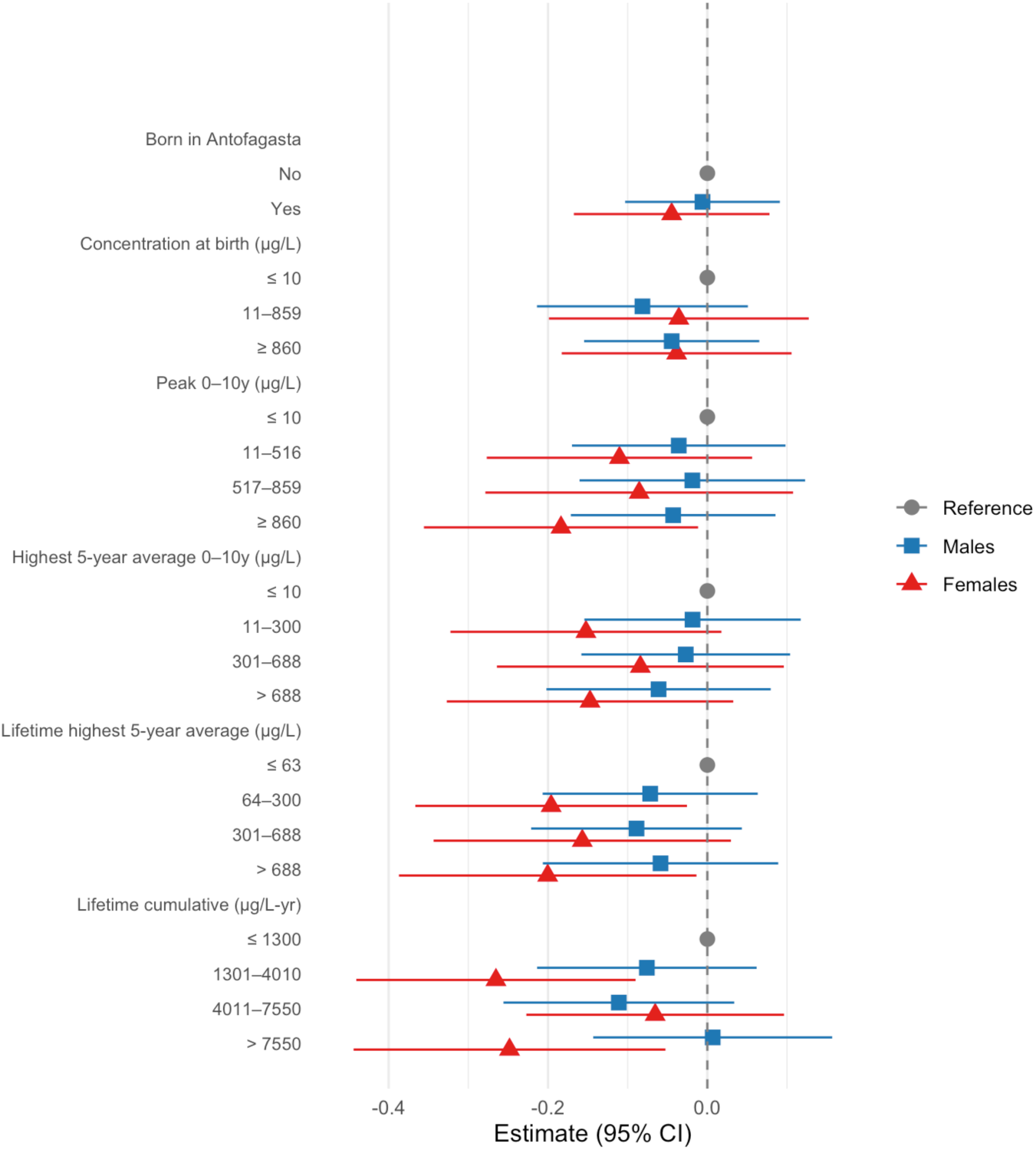
Forrest plot of regression estimates (mean difference with 95% CI) for log-transformed cortisol concentrations associated with arsenic exposure metrics stratified by sex. Exposure measures include being born in Antofagasta; arsenic concentration at birth (μg/L); peak exposure from 0 to 10 years (μg/L); highest 5-year average from 0 to 10 years (μg/L); lifetime highest 5- year average (μg/L); and lifetime cumulative exposure (μg/L-yr). Reference groups are in grey (circle) with effect estimates for males in blue (square) and females in red (triangle).

Lastly, we conducted stratified analyses to evaluate potential confounding by age and recruitment period. When stratifying results by median age, the directions of associations between arsenic exposure metrics and plasma cortisol concentrations was generally consistent for participants ≤ 52 years and >52 years **(Supplemental Table 3**). One exception was that statistically significant associations were observed only among participants ≤52 years for the highest quartile of highest 5-year average exposure during ages 0–10 years, the second and fourth quartiles of lifetime highest 5-year average exposure, and the second quartile of cumulative lifetime exposure. Tests for interaction by age (≤52 vs >52 years) were not statistically significant for any exposure metric. We also examined differences by recruitment period. Associations between arsenic exposure metrics and cortisol concentrations were generally more negative among participants recruited in 2013 compared to those recruited in 2017 (**Supplemental Table 4**). Statistically significant associations were only observed among participants recruited in 2013 and for the highest quartiles of peak exposure before the age of 10 years, 5-year average before the age of 10 years, and lifetime highest 5-year average. Tests for interaction by recruitment year were not statistically significant.

## 4. Discussion

This study provides the first evidence that early-life arsenic exposure may be associated with lower plasma cortisol concentrations in adults. These effects were observed over 40 years after exposure cessation among individuals currently living in Antofagasta, Chile—a city with a well-documented distinct period of past arsenic exposure. We also observed that associations between higher arsenic exposure and lower cortisol concentrations were stronger among female participants, indicating effect modification by sex. Another original aspect of this work is the application of a novel low-cost and rapid cell-based assay to measure cortisol equivalent concentrations in plasma for environmental epidemiology studies. Overall, this work demonstrates that arsenic exposure may have persistent endocrine-disrupting effects that alter cortisol levels in adulthood.

Observed correlations between sociodemographic factors and plasma cortisol equivalent concentrations were consistent with previous studies. Plasma cortisol equivalent concentrations in the study population ranged from 2.57-14.43 μg/dL, with a median of 6 μg/dL. These values fell within serum cortisol reference ranges of 7-25 μg/dL in the morning and 2-14 μg/dL in the evening (Mayo Clinic Laboratories, 2025). When using spline regression to examine the relationship between time of blood draw and plasma cortisol equivalent concentrations, we observed that cortisol levels were highest among participants with collection times between 6-8AM and lower levels were observed with later collection times. This result aligns with the diurnal cycle of cortisol, which typically peaks 30 minutes after waking and gradually declines throughout the day (Oster et al., 2017). While major limitations of this study were that we did not record waking time or conduct repeat cortisol sampling to assess individual diurnal patterns, we observed the clinically expected diurnal peak at 6-8AM at the population level (Mayo Clinic Laboratories, 2025). Since samples were collected using the same procedures in all participants, regardless of past arsenic exposure, the resulting bias would most likely be towards the null.

Additionally, our results are likely not confounded by time of collection since there was no difference between arsenic exposure groups. Future studies should collect multiple samples throughout the day to better evaluate the effect of arsenic exposure on diurnal cortisol secretion patterns. Additionally, we observed that morning plasma cortisol concentrations were lower among female participants than males. This sex difference in cortisol is consistent with previous studies that reported men had higher levels of morning serum cortisol (Klinger-König et al., 2021) and hair cortisol concentrations (Stalder et al., 2017) than women. Estrogen administration has been shown to decrease morning cortisol levels, suggesting that these differences between males and females may have been driven by sex hormones (Edwards and Mills, 2008). Our findings are generally in line with expected physiological patterns, strengthening the reliability of our cortisol measurements.

We used a unique study population from Northern Chile with high arsenic exposure during childhood, which may limit generalizability of our findings. However, investigating associations in populations where exposures are high, at least initially, can provide insights into the direction and magnitude of the relationship between arsenic exposure and circulating cortisol levels. We observed that individuals with a peak lifetime 5-year average exposure to arsenic above 63 μg/L had significantly lower morning plasma cortisol concentrations compared to those with lower arsenic exposure, with similar trends observed for high exposure during ages 0-10 years (both peak and 5-year average). While only the association with peak lifetime 5-year average arsenic exposure was statistically significant, effect estimates were similar to those for peak 5-year average arsenic exposure between age 0-10y (p =0.29). Although the bulk of exposure occurred during between ages 0-10 years, these results indicate that exposure to arsenic after age 10y may also play a role in influencing cortisol regulation later in life. Moreover, when assessing lifetime cumulative arsenic exposure, individuals in the second and fourth exposure quartiles had lower morning plasma cortisol concentrations compared to those in the first exposure quartile, with a greater difference observed for the second quartile. This pattern may suggest a non-linear relationship between cumulative arsenic exposure and cortisol concentrations, although interpretation is limited by the small sample size. Our results are consistent with the only other study that examined the relationship between early-life arsenic exposure and cortisol levels. A study of infants conducted in Arica, a city in Chile located approximately 700km from our study population, observed lower salivary cortisol levels among high income children in the highest tertile of arsenic exposure during the second trimester of pregnancy (Salgado et al., 2019). Our findings build on this work by demonstrating that early-life arsenic exposure has persistent effects on cortisol levels later in life. Our study population also had generally high socioeconomic status (assessed via self-reported material items), making our study comparable to the Arica study in terms of geography, exposure timing, and socioeconomic status. However, important methodological differences limit direct comparison between the two studies. The Arica study assessed exposure by measuring total urinary inorganic arsenic (the sum of As^V^, As^III^, MMA, and DMA), which reflects recent exposure from multiple potential sources. Reported arsenic exposure levels in Arica (2.05−69.3 μg/L) were substantially lower than those in our study (0−860 μg/L), consistent with historically lower concentrations of arsenic in Arica’s drinking water (Ferreccio et al., 2013). Therefore, urine measurements in Salgado et al. likely reflected other sources such as soil or secondary air pollution. Additionally, the study in Arica measured salivary cortisol in children, which captures free bioactive hormone, whereas we measured plasma cortisol in adults that reflects total circulating cortisol. Despite these methodological differences, the consistent observation of lower cortisol associated with early-life arsenic exposure in both studies strengthens our findings. Lower morning GC levels have been previously linked to both early-life exposures (e.g. smoking, lead, childhood adversity) and higher levels of cardiovascular risk factors including blood pressure (Stroud et al., 2014; Braun et al., 2014; Power et al., 2012; Kuras et al., 2017; Rosmond and Björntorp, 2000). Consequently, associations between early-life exposures, such as arsenic, and chronic disease risk in adulthood may be mediated in part by reductions in cortisol levels.

In contrast to our results, a study conducted among women from West Bengal found that low-level urinary arsenic exposure (11-50 μg/L) was associated with a two-fold increase in serum cortisol (Sinha et al., 2014). However, our study population experienced substantially higher levels of arsenic exposure compared to West Bengal (860 vs. 50 μg/L). In addition, the exposure window for our study population differs from the West Bengal study. The exposure levels in West Bengal were similar to concentrations tested in animal studies, where prenatal exposure was associated with elevated plasma corticosterone in adulthood (Martinez et al., 2008; Goggin et al., 2012). *In vitro* arsenic has a biphasic effect on the glucocorticoid receptor with lower concentrations inducing activation and higher concentrations having inhibitory effects, which might explain differences observed between studies with high versus low exposure levels (Bodwell et al., 2006). Future studies should examine whether varying levels of arsenic exposure exert differential effects on the HPA axis. Null associations were also reported in other studies conducted among pregnant women and adult residents near e-waste recycling facilities in China (Chen et al., 2024; Li et al., 2020). These studies may have been confounded by the reliance on plasma arsenic concentrations, which only reflects recent exposure due to its short half-time in humans (ATSDR, 2007). We also observed null associations when arsenic exposure was assessed using place of birth and concentration at birth. However, the highest 5-year average (both lifetime and between age 0-10 years) and lifetime cumulative exposure metrics that reflect both intensity and duration of exposure were associated with lower plasma cortisol levels. These findings align with what is known about HPA axis development, which begins during gestation and continues throughout puberty (Howland et al., 2017). Arsenic exposures during this extended developmental window may lead to persistent changes to HPA axis regulation. For example, mice exposed to 50ppb arsenic from birth through weaning had elevated plasma corticosterone levels at post-natal day 35 (Goggin et al., 2012), whereas mice exposure only prenatally (until birth) showed reduced corticosterone at postnatal day 60 (Sobolewski et al., 2018). These studies highlight that both the timing and duration of arsenic exposure may significantly influence its endocrine disrupting effects. Our findings provide human evidence that chronic early-life arsenic exposure, particularly before age 10, may alter cortisol secretion patterns in adulthood. Prospective longitudinal studies are needed to characterize the exposure-response relationship and identify critical windows of susceptibility.

The observation of lower plasma cortisol concentrations 40-50 years after exposure ended suggests that arsenic has a persistent effect on HPA axis function. This lasting effect may be mediated by epigenetic mechanisms, such as DNA methylation of the glucocorticoid receptor (GR). Studies have shown that epigenetic marks set by early-life exposures at the GR gene locus (*NR3C1*) remain stable throughout life (McGowan et al., 2009; Radtke et al., 2011).

Epidemiological studies have reported that prenatal arsenic exposure is associated with increased methylation of *NR3C1* in placental tissues (Appleton et al., 2017; Cardenas et al., 2015).

Additionally, trophoblasts treated with arsenic *in vitro* exhibited dose-dependent changes in DNA methylation at CpG sites across 12 genes involved in GR signaling, which corresponded with altered gene expression (Meakin et al., 2019). These findings suggest that early-life arsenic exposure may lead to lasting disruption in glucocorticoid signaling and HPA axis regulation through DNA methylation changes, though further studies are needed to confirm this mechanism.

Associations between arsenic exposure and lower plasma cortisol concentrations were more pronounced among female participants, suggesting potential sex-specific biological effects. Specifically, women in the second and fourth quartiles of lifetime cumulative exposure had approximately 23.3% and 22.0% lower cortisol concentrations, respectively, compared with women in the lowest exposure quartile. In contrast, the cortisol differential among males was minimal and did not reach statistical significance at any level of cumulative exposure. These results align with prior animal studies demonstrating that developmental arsenic exposure can induce sex-specific changes in gene expression related to HPA axis signaling (Allan et al., 2015; Solomon et al., 2020). For example, corticotropin-releasing hormone (CRH) mRNA levels were elevated in the frontal cortex of arsenic-exposed female mice at postnatal day 70 but reduced in exposed male mice (Solomon et al., 2020). CRH is produced by the hypothalamus and initiates the stress response by stimulating ACTH release from the pituitary gland, which in turn promotes cortisol secretion from the adrenal gland. Individuals with post-traumatic stress disorder often exhibit elevated CRH with lower cortisol levels, suggesting that arsenic may induce a similar dysregulation of the HPA axis (Raglan et al., 2017). Normally, glucocorticoids suppress CRH transcription through negative feedback mechanisms and removal of endogenous glucocorticoids increase CRH gene expression (Evans et al., 2013). Arsenic exposure in females may increase CRH levels, which disrupts this feedback mechanism and may explain the more pronounced impairment of HPA axis regulation. In addition to CRH, arsenic exposure influences expression of other genes involved in glucocorticoid signaling in a sex-specific manner. In male mice, developmental exposure altered *FKBP1* and *HSD11B1* expression in ways that would reduce sensitivity to glucocorticoids, potentially reflecting a compensatory mechanism not overserved in females (Solomon et al., 2020). The stronger associations observed among female participants in our study may reflect a lack of these adaptive responses. Another possible mechanism underlying the more pronounced effects observed among females could involve modulation by sex hormones, such as estradiol. While estradiol was not measured in this study, arsenic exposure has been shown to increase aromatase expression and elevate estradiol confrontations in human breast epithelial cells (Xu et al., 2014). Studies have also demonstrated that estradiol impairs glucocorticoid-dependent negative feedback of the HPA axis (Weiser and Handa, 2009). Therefore, if arsenic increases estradiol levels in females, it might explain the altered cortisol concentrations observed among those with high arsenic exposure. The relationship between early-life arsenic, estradiol concentrations, and HPA axis regulation warrants further investigation.

Cortisol equivalent concentrations were measured using a rapid and low-cost cell-based bioassay, which is not specific to cortisol since it quantifies total activation of the glucocorticoid receptor induced by all compounds present in plasma. In a study of 12 healthy individuals, serum cortisol concentrations measured using this GR bioassay were highly correlated with values obtained from a cortisol enzyme-linked immunosorbent assay (de la Rosa et al., 2021).

Therefore, assay activity in the current study likely reflects plasma cortisol concentrations but should be confirmed using analytical techniques that specifically measure cortisol (e.g. ELISA or mass spectrometry), as other compounds present in plasma that either activate or inhibit GR could potentially influence assay results and cause deviations from true plasma cortisol concentrations. For instance, arsenic and its methylated metabolites have been shown to inhibit GR signaling *in vitro* at concentrations as low as 1µM (∼75 µg/L) (Gosse et al., 2014; Bodwell et al., 2006, 2004; Kaltreider et al., 2001). However, current arsenic exposure is unlikely to explain the observed reductions in cortisol equivalent plasma concentrations associated with arsenic exposures that occurred decades before sample collection. Furthermore, annual arsenic levels in drinking water at the time of sample collection were at or below 10 µg/L (∼133.5 nM) for all participants, which is nearly tenfold lower than the concentrations shown to inhibit GR activity *in vitro*.

A limitation of this study was the relatively small sample size. Many samples are needed to detect changes in cortisol levels given the high amount of inter-individual variability (Almeida et al., 2009). To address this, we combined samples collected in 2013 and 2017. Combining samples from these two collection periods could potentially increase variability and heterogeneity because of temporal and/or batch effects, consequently reducing our ability to detect differences. We attempted to mitigate these issues by using consistent sample processing protocols across both recruitment periods and normalization between batches using a common reference sample included on ever plate. Furthermore, results were similar in a sensitivity analysis stratified by recruitment period. Despite the limited sample size, the statistically significant associations observed indicate that our study possessed adequate power to detect a difference in morning plasma cortisol concentrations with arsenic exposure. Associations between early-life arsenic exposure and plasma cortisol levels should be further investigated in a larger study.

Confounding is also possible. Adjustments for current smoking status, education level, chronic medical conditions (hypertension, diabetes, or cancer), and technical factors (plate and recruitment period) had little impact on our results. While inaccurate recall of residential history could have led to exposure misclassification, this is unlikely given that individuals generally have reliable knowledge of where they have lived. Arsenic exposure may also occur through dietary sources, such as rice and rice products, apple juice, beer and wine, and seafood (Nachman et al., 2017). However, most food in this population is imported from areas with low arsenic water concentrations since climate in the study area is so dry. It is also unlikely that our findings with cumulative arsenic exposure are confounded by age. Plasma cortisol concentrations were not correlated with age and effect sizes changed very little with adjustment for age.

Moreover, the direction of associations remained consistent in age-stratified analyses, suggesting that age is not a driver of observed associations.

Studies examining the relationship between environmental exposures and cortisol levels in humans remain limited. Our findings provide epidemiological evidence of arsenic’s endocrine disrupting effects. The adverse effects of arsenic on endocrine-related endpoints, such as cortisol levels, should be investigated further and considered when establishing regulatory standards.

Future studies should also assess whether cortisol mediates the association between arsenic exposure and disease.

## Data Availability

All data produced in the present study are available upon reasonable request to the authors

## Acknowledgements

This project was supported by NIEHS grants R01ES014032, P42ES004705, K99/R00ES024808, and R21ES035517. The views expressed in this paper are solely those of the authors and do not necessarily reflect those of NIEHS. We thank Jen-Chywan Wang, Andres Cardenas, Felicia Castriota, and Rachael Philips for their consultation and discussion of our findings.

