## Supplemental Tables for "Long-Term Effect of Early-Life Arsenic Exposure on Morning Plasma Cortisol in Adults from Antofagasta, Chile"

| Supplemental Table 1. Sociodemographic variables by recruitment period | | | |
| --- | --- | --- | --- |
| **Variable** | **2013 Study (N=84)** | **2017 Study (N=148)** | **p-value** |
| **Age** (years), median (IQR) | 48.0 (45.0, 52.0) | 54.0 (50.0, 56.0) | <0.001 |
| **Sex,** n (%) |  |  |  |
| Male | 54 (64.3) | 74 (50.0) | 0.04 |
| Female | 30 (35.7) | 74 (50.0) |  |
| **Smoking Status**, n (%) |  |  | 0.10 |
| Never | 28 (33.3) | 66 (44.6) |  |
| Ever | 56 (66.7) | 82 (55.4) |  |
| **Education**, n (%) |  |  |  |
| Completed secondary or higher | 74 (88.1) | 112 (75.7) | 0.02 |
| Less than secondary | 9 (10.7) | 35 (23.6) |  |
| Missing | 1 (1.2) | 1 (0.7) |  |
| **Blood Collection Time** (AM), median (IQR) | 9:05 (7:57, 10:06) | 8:50 (8:30, 9:40) | 0.92 |
| **Arsenic Exposure Metrics,** median (IQR) |  |  |  |
| Water concentration at birth (μg/L) | 130.0 (6.2, 860) | 250 (0.75, 860) | 0.43 |
| Peak exposure from birth to 10 years (µg/L) | 250 (6.2, 688) | 662 (130, 860) | 0.05 |
| Highest 5-year average from birth to 10 years (µg/L) | 190.1 (6.2, 602.0) | 430 (6.2, 602) | 0.02 |
| Lifetime highest 5-year average (μg/L) | 215 (38.1, 602) | 430 (119, 774) | 0.002 |
| Lifetime cumulative (μg/L-yr) | 2688.2 (773.6, 5489.1) | 4736.7 (2180.8, 8557) | 0.003 |
| Lifetime average after age 20 (μg/L) | 26.0 (15.4, 30.1) | 24.8 (17.8, 30.4) | 0.73 |

| Supplemental Table 2. Associations between arsenic exposure and plasma cortisol equivalent concentrations by sex | | | | |
| --- | --- | --- | --- | --- |
| **Arsenic Exposure Metric** | **Males ß (95% CI)^1^** |  | **Females ß (95% CI)^1^** | **p-interaction** |
| **Born in Antofagasta** |  |  |  | 0.53 |
| No | Ref |  | Ref |  |
| Yes | -0.006 (-0.103, 0.091) |  | -0.045 (-0.168, 0.078) |  |
| **Concentration at birth (μg/L)** |  |  |  | 0.84 |
| ≤ 10 | Ref |  | Ref |  |
| 11-859 | -0.081 (-0.214, 0.051) |  | -0.036 (-0.199, 0.127) |  |
| 860 | -0.045 (-0.155, 0.065) |  | -0.039 (-0.183, 0.106) |  |
| **Peak Exposure 0-10 years (μg/L)** |  |  |  | 0.34 |
| ≤ 10 | Ref |  | Ref |  |
| 11-516 | -0.036 (-0.170, 0.098) |  | -0.110 (-0.277, 0.056) |  |
| 517-859 | -0.019 (-0.160, 0.123) |  | -0.086 (-0.279, 0.108) |  |
| 860 | -0.043 (-0.171, 0.086) |  | -0.184 (-0.356, -0.012) |  |
| **Highest 5-year average from** **0-10 years (μg/L)** |  |  |  | 0.36 |
| ≤ 10 | Ref |  | Ref |  |
| 11-300 | -0.019 (-0.154, 0.117) |  | -0.152 (-0.323, 0.018) |  |
| 301-688 | -0.027 (-0.158, 0.104) |  | -0.084 (-0.264, 0.096) |  |
| ≥ 689 | -0.061 (-0.202, 0.080) |  | -0.147 (-0.327, 0.033) |  |
| **Lifetime highest 5-year average (μg/L)** |  |  |  | 0.26 |
| ≤ 63 | Ref |  | Ref |  |
| 64-300 | -0.072 (-0.207, 0.063) |  | -0.196 (-0.367, -0.026) |  |
| 301-688 | -0.089 (-0.221, 0.043) |  | -0.157 (-0.344, 0.030) |  |
| ≥ 689 | -0.059 (-0.207, 0.089) |  | -0.200 (-0.387, -0.014) |  |
| **Lifetime cumulative (μg/L-yr)** |  |  |  | 0.04 |
| ≤ 1300 | Ref |  | Ref |  |
| 1301-4010 | -0.076 (-0.214, 0.062) |  | -0.265 (-0.441, -0.090) |  |
| 4011-7550 | -0.111 (-0.256, 0.034) |  | -0.065 (-0.227, 0.096) |  |
| ≥ 7551 | 0.007 (-0.143, 0.156) |  | -0.248 (-0.444, 0.053) |  |
| 1. Models are adjusted for age (continuous), sex, education (< secondary vs. ≥ secondary), smoking status (ever vs. never),and time of blood collection (hours since 6AM, modeled as a spline with 3 degrees of freedom) | | | | |

| Supplemental Table 3. Associations between arsenic exposure and plasma cortisol equivalent concentrations by age group | | | | |
| --- | --- | --- | --- | --- |
| **Arsenic Exposure Metric** | **≤52 years old ß (95% CI)^1^** |  | **>52 years old ß (95% CI)^1^** | **p-interaction** |
| **Born in Antofagasta** |  |  |  | 0.19 |
| No | Ref |  | Ref |  |
| Yes | 0.026 (-0.080, 0.132) |  | -0.063 (-0.180, 0.053) |  |
| **Concentration at birth (μg/L)** |  |  |  | 0.90 |
| ≤ 10 | Ref |  | Ref |  |
| 11-859 | -0.064 (-0.206, 0.078) |  | -0.031 (-0.195, 0.132) |  |
| 860 | -0.025 (-0.145, 0.095) |  | -0.038 (-0.176, 0.101) |  |
| **Peak Exposure 0-10 years (μg/L)** |  |  |  | 0.95 |
| ≤ 10 | Ref |  | Ref |  |
| 11-516 | -0.088 (-0.226, 0.051) |  | -0.041 (-0.220, 0.139) |  |
| 517-859 | -0.061 (-0.222, 0.101) |  | -0.029 (-0.203, 0.145) |  |
| 860 | -0.124 (-0.266, 0.017) |  | -0.056 (-0.222, 0.111) |  |
| **Highest 5-year average from** **0-10 years (μg/L)** |  |  |  | 0.69 |
| ≤ 10 | Ref |  | Ref |  |
| 11-300 | -0.090 (-0.224, 0.044) |  | -0.074 (-0.281, 0.132) |  |
| 301-688 | -0.040 (-0.187, 0.108) |  | -0.052 (-0.219, 0.114) |  |
| > 688 | -0.174 (-0.335, -0.014) |  | -0.022 (-0.185, 0.141) |  |
| **Lifetime highest 5-year average (μg/L)** |  |  |  | 0.72 |
| ≤ 63 | Ref |  | Ref |  |
| 64-300 | -0.156 (-0.289, -0.023) |  | -0.058 (-0.252, 0.137) |  |
| 301-688 | -0.104 (-0.251, 0.042) |  | -0.107 (-0.279, 0.065) |  |
| > 688 | -0.173 (-0.335, -0.012) |  | -0.049 (-0.219, 0.121) |  |
| **Lifetime cumulative (μg/L-yr)** |  |  |  | 0.65 |
| ≤ 1300 | Ref |  | Ref |  |
| 1301-4010 | -0.171 (-0.309, -0.034) |  | -0.162 (-0.364, 0.040) |  |
| 4011-7550 | -0.106 (-0.243, 0.032) |  | 0.018 (-0.169, 0.204) |  |
| > 7550 | -0.138 (-0.365, 0.090) |  | -0.063 (-0.228, 0.102) |  |
| 1. Models are adjusted for sex, education (< secondary vs. ≥ secondary), smoking status (ever vs. never), recruitment year, and time of blood collection (hours since 6AM, modeled as a spline with 3 degrees of freedom) | | | | |

| Supplemental Table 4. Associations between arsenic exposure and plasma cortisol equivalent concentrations by recruitment period | | | | |
| --- | --- | --- | --- | --- |
| **Arsenic Exposure Metric** | **2013 Study ß (95% CI)^1^** |  | **2017 Study ß (95% CI)^1^** | **p-interaction** |
| **Born in Antofagasta** |  |  |  | 0.30 |
| No | Ref |  | Ref |  |
| Yes | 0.034 (-0.091, 0.160) |  | -0.041 (-0.142, 0.059) |  |
| **Concentration at birth (μg/L)** |  |  |  | 0.91 |
| ≤ 10 | Ref |  | Ref |  |
| 11-859 | -0.007 (-0.178, 0.163) |  | -0.057 (-0.193, 0.079) |  |
| 860 | -0.038 (-0.179, 0.102) |  | -0.031 (-0.149, 0.086) |  |
| **Peak Exposure 0-10 years (μg/L)** |  |  |  | 0.73 |
| ≤ 10 | Ref |  | Ref |  |
| 11-516 | -0.050 (-0.207, 0.107) |  | -0.069 (-0.214, 0.076) |  |
| 517-859 | -0.055 (-0.234, 0.124) |  | -0.056 (-0.207, 0.096) |  |
| 860 | -0.185 (-0.349, -0.021) |  | -0.050 (-0.189, 0.090) |  |
| **Highest 5-year average from** **0-10 years (μg/L)** |  |  |  | 0.75 |
| ≤ 10 | Ref |  | Ref |  |
| 11-300 | -0.073 (-0.230, 0.083) |  | -0.072 (-0.221, 0.078) |  |
| 301-688 | -0.058 (-0.224, 0.108) |  | -0.056 (-0.201, 0.089) |  |
| ≥ 689 | -0.202 (-0.401, -0.004) |  | -0.046 (-0.191, 0.099) |  |
| **Lifetime highest 5-year average (μg/L)** |  |  |  | 0.36 |
| ≤ 63 | Ref |  | Ref |  |
| 64-300 | -0.205 (-0.364, -0.047) |  | -0.074 (-0.222, 0.074) |  |
| 301-688 | -0.129 (-0.291, 0.034) |  | -0.111 (-0.260, 0.037) |  |
| ≥ 689 | -0.270 (-0.466, -0.073) |  | -0.047 (-0.197, 0.104) |  |
| **Lifetime cumulative (μg/L-yr)** |  |  |  | 0.65 |
| ≤ 1300 | Ref |  | Ref |  |
| 1301-4010 | -0.240 (-0.412, -0.069) |  | -0.133 (-0.282, 0.016) |  |
| 4011-7550 | -0.046 (-0.206, 0.114) |  | -0.058 (-0.208, 0.016) |  |
| ≥ 7551 | -0.267 (-0.456, 0.114) |  | -0.037 (-0.198, 0.123) |  |
| 2. Models are adjusted for age (continuous), sex, education (< secondary vs. ≥ secondary), smoking status (ever vs. never),and time of blood collection (hours since 6AM, modeled as a spline with 3 degrees of freedom) | | | | |

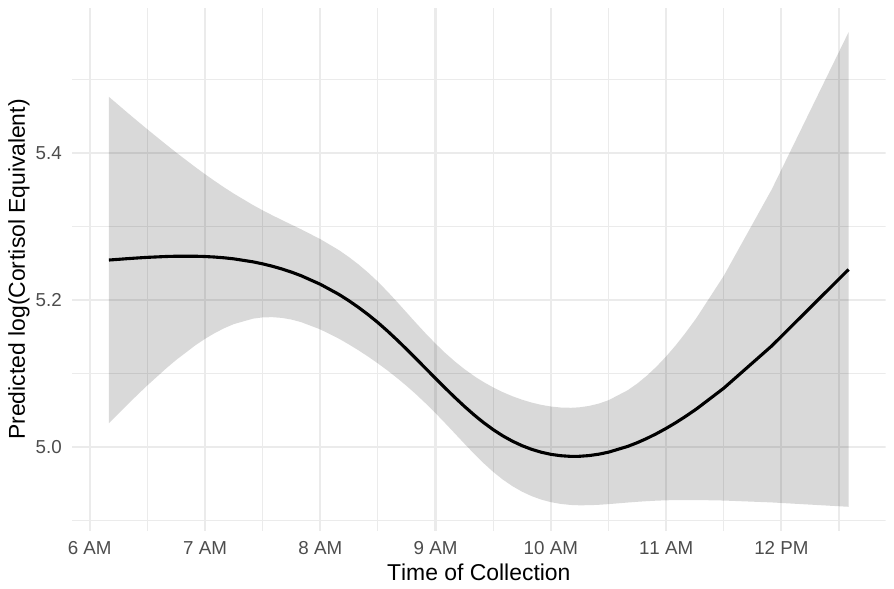

**Supplemental Figure 1.** Predicted plasma cortisol equivalent concentration (log transformed) from 6:10 AM to 12:35 PM with collection time modeled as a nature spline with 3 degrees of freedom. The curve illustrates the peak cortisol levels occurred around 7:00AM, with a sharp decline occurring thereafter until approximately 10:00 AM, followed by a stabilization through the early afternoon.

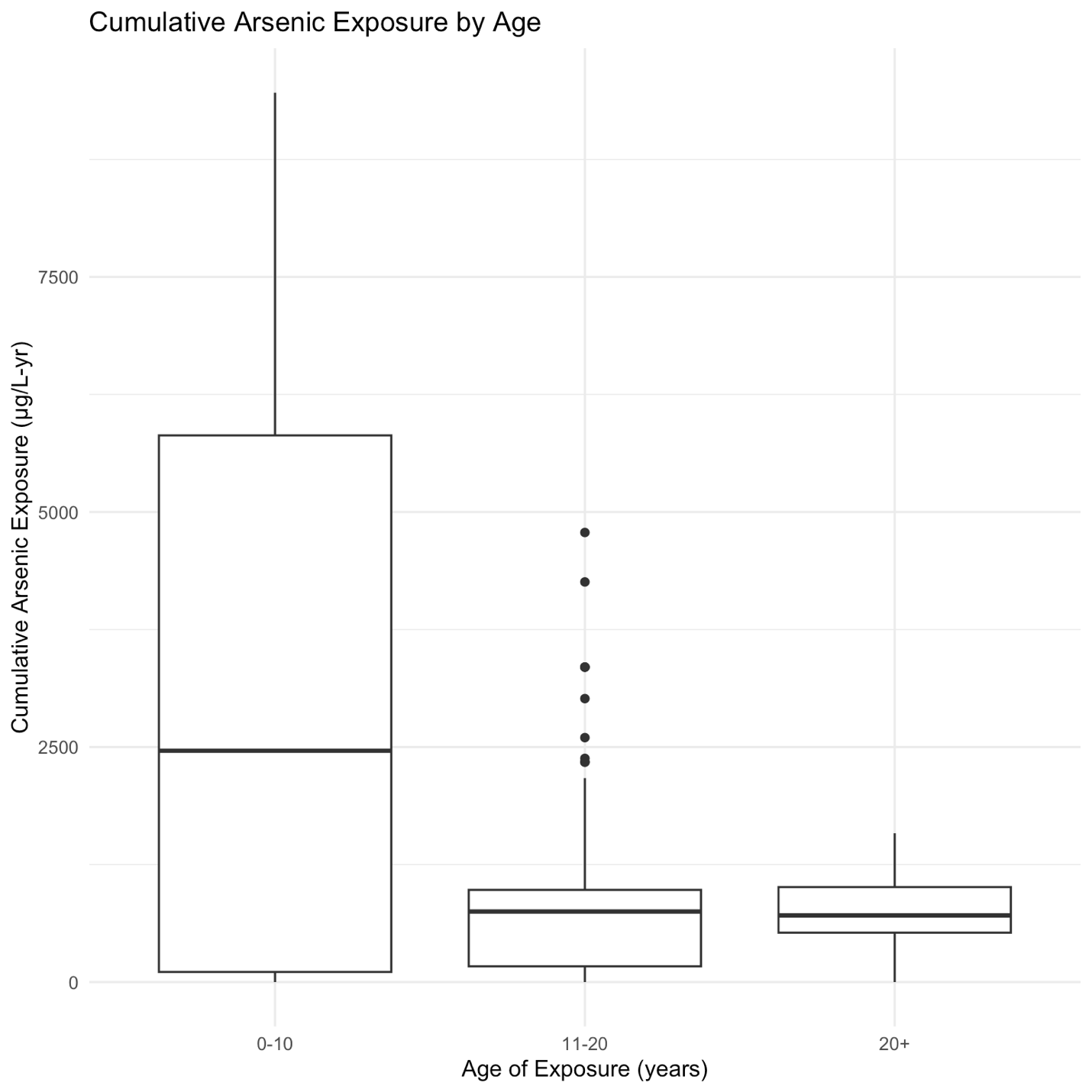

**Supplemental Figure 2.** Cumulative arsenic exposure by age. Boxplot of cumulative exposure (ug/L-years) that occurred during the ages of 0-10, 11-20, and >20 years old.
